# Meperidine’s Continued Fall and Regional Variance (2019-2023): Associations with Income and Obesity

**DOI:** 10.1101/2025.07.16.25331668

**Authors:** Brandon Gonzalez, Lauren G. Healy, Lavinia R. Harrison, Brian J. Piper

## Abstract

**Objectives:** Meperidine, once viewed as relatively safe, is now discouraged in clinical settings due to its associated risks. Previous studies have identified a significant decrease in meperidine distribution across the US from 2000 to 2021. Regional disparities accompanied this decline. The goal of this study was to investigate if the decrease in meperidine distribution has continued in recent years: 2019 to 2023; and if regional variations persist. This investigation also aimed to identify correlates of meperidine distribution, including adult obesity prevalence and annual income, to provide insight into the regional variation.

**Design:** Retrospective observational study utilizing data from the Automation of Reports and Consolidated Orders System (ARCOS) Drug Retail Summary Reports by the Drug Enforcement Administration (DEA), the CDC, and the US Census Bureau.

**Setting:** USA, including Puerto Rico

**Participants:** US population

**Primary and secondary outcome measures:** The primary outcome was the meperidine distribution across the US between 2019 and 2023. Secondary outcomes included associations between meperidine distribution and adult obesity prevalence and median household income.

**Results:** Total meperidine distribution across the US dropped by 57.8% from 2019 to 2023. A substantial geographic variation was found with southern states accounting for the 2nd, 3rd, and 4th highest in meperidine use capita in 2023, only behind Puerto Rico. In contrast, northeastern states accounted for four of the five lowest states. A significant relationship was found between annual income and meperidine distribution in 2022 (r(49) = -.38, p < .005). A trend was observed between adult obesity prevalence and meperidine distribution (r(52) = +.24, p = .078).

**Conclusion:** Our study revealed a continued decrease in meperidine distribution and continued presence of geographical variation from 2019 to 2023. Furthermore, a novel relationship was identified between meperidine distribution and annual income.

**Strengths and Weaknesses:** Strengths

- The ARCOS database is comprehensive for meperidine distribution in every year/location used in this study.
- This study reports on the pronounced state level differences in meperidine use and the association with other variables which include annual income and adult obesity prevalence by location.

Weaknesses

- The Automation of Reports and Consolidated Orders System database lacks information on formulations and the number of prescriptions.
- Further study should examine spending on meperidine by Medicaid, Medicare, or private insurance agencies.

## INTRODUCTION

Meperidine was once the most used opioid in the US, but has since been classified as a dangerous analgesic.^1, 2^ Due to initial studies in the mid 20th century, it was believed that meperidine had advantages over the standard at the time, morphine.^2^ However, the identification of multiple risk factors associated with meperidine led to a change in perspective.^2^ The use of meperidine carries the risks of serotonin syndrome, elevated concentration of neurotoxic normeperidine, and the potential for misuse due to its pharmacokinetics.^3^ Prior papers have discussed the tragic case of Libby Zion, in which a young patient died due to an interaction between meperidine and the antidepressant, phenelzine, which led to a fatal serotonin syndrome. This event would not only lead to national changes in terms of medical education, but also to a reexamination for the use of meperidine in clinical settings.^3,4^ Other research would also discover that taking several different Serotonin-Norepinephrine reuptake inhibitors with meperidine also risked causing serotonin syndrome.^5^ The US Drug Enforcement Administration (DEA), classified meperidine as a Schedule II controlled substance due to it’s high potential for abuse and addiction since 1970, a decision which persists today. ^6,7^ Furthermore, these findings supported the World Health Organization’s (WHO) decision to remove meperidine from their *List of Essential Medicines* in 2003.^3^ In addition, the 2019 *American Geriatrics Society Beers Criteria* has recommended to avoid meperidine with the strength of the recommendation as “Strong”.^8^

Prior studies have identified significant decreases in meperidine distribution in the US between 2000 and 2021.^3,4^ While these investigations revealed reductions in every US state, substantial geographic variations were also observed. Southern states such as Mississippi, Alabama, Arkansas, and Oklahoma, had significantly higher rates of meperidine distribution when compared to the nation as a whole, and more specifically, the northeast.^3,4^ Past papers, in an effort to further understand this geographic variation of meperidine distribution, attempted to find an association between meperidine distribution and adult obesity prevalence per state. Results indicated a significant relationship between the two variables.^3^ Altogether, these reports indicate that meperidine use is significantly decreasing in the US with substantial geographic variation that may be explained by different variables, such as adult obesity prevalence.^3,4^

While these studies offer valuable insight into the changes of meperidine distribution from 2000 to 2021, there is a need for data on more recent usage patterns.^3,4^ Furthermore, additional investigation into possible correlates of meperidine distribution across the US is also valuable in order to gain more insight into the sizeable variations by state. This report answers three questions. First, whether there was continued decreases in meperidine distribution from 2019 to 2023. Second, if the presence of substantial regional variations persist. Third, if adult obesity prevalence and/or annual income by region are correlates that can offer insight into the variations in meperidine distribution by state. We hypothesized that there would be a continued decrease in meperidine usage with significant correlations from obesity and annual income by region.

## METHODS

### Procedure

Data on meperidine distribution across the 50 States, the District of Columbia, the Virgin Islands, Puerto Rico, and Guam were collected via the 2019 to 2023 ARCOS Drug Retail Summary Reports as published by the DEA.^9^ Meperidine distribution for the first half of 2024, the most recent available, was also collected from ARCOS.^9^ Data was collected from the Report 2 and Report 7 sections, which included information on drug distribution by state and retail drug purchases. Populations for the 50 US states and Puerto Rico were found within the American Community Survey’s 1 - Year Estimates Data Profiles, as published by the US Census Bureau, for the years 2019, 2021, and 2022.^10^ Table 2 of the 2020 US Census, which included resident population numbers, was utilized to find the population of the 50 States and Puerto Rico for 2020.^11^ The 2023 populations for the 50 states and Puerto Rico were obtained from the National Population Totals and Components of Change, also published by the US Census Bureau.^12^ The United Nations Data Portal Population Division online tool was used to find the populations of the Virgin Islands and Guam for 2019 to 2023.^13^ Populations for the 50 US states and Puerto Rico for the first half of 2024 were collected from the United States Census Bureau Vintage 2024 data.^14^ The percentage of adults with obesity for the 50 States and Puerto Rico for 2022 was collected from the Nutrition, Physical Activity, and Obesity: Data, Trends and Maps online data tool as published by the CDC.^15^ Median household incomes per state for 2022 and 2023 were found in the American Community Survey Briefs: Household Income in States and Metropolitan Areas: 2022 and 2023.^16, 17^ P values, regression statistics, and graphs were generated via GraphPad Prism 10.4.2, San Diego, CA. All heat maps were generated via Datawrapper, Berlin, Germany. Heat maps showing meperidine distribution by category were generated for the states with the highest meperidine distribution in 2023. Procedures were approved as exempt by the Geisinger IRB.

## DATA ANALYSIS

Milligrams of meperidine use per 100 individuals for the years 2019 to 2023 were calculated in IBM SPSS Statistics by using the meperidine usage data found in Report 2 of the ARCOS Drug Retail Summary Reports and the population data found in both the American Community Survey and U.S.Census.^9, 10, 11, 12^ States were then organized in descending order of the resultant meperidine usage for the year 2023 in a waterfall plot with significance being defined as values above 1.96 standard deviations from the mean. A heatmap of meperidine usage per 100k individuals in the 50 states and Puerto Rico for 2023 was generated in order to visualize this data. An additional waterfall plot containing the percent reduction of meperidine use per 100 individuals between the years 2019-2023 was generated as well. Linear regressions were then generated to find associations between meperidine usage with median household income in 2022 and 2023 and adult obesity prevalence per region for 2022. Figures and analyses were completed via GraphPad Prism 10, San Diego, CA.

## RESULTS

The total distribution of meperidine decreased from 292,694,240 milligrams (mg) in 2019 to 123,433,330 mg in 2023, a 57.8% reduction.^9^ In the same four-year period, the mean meperidine use in mg per 100 individuals declined from 98.3 to 37.2, a 62.2% decrease. While the median meperidine use in mg per 100 individuals diminished from 79.9 to 25.3, a 68.3% decrease (Figure 1). The waterfall plot, which visualized meperidine usage in mg per 100 individuals in 2023, revealed significance in one state and one US territory (Figure 2). Puerto Rico (193.5) and Arkansas (141.7) were significantly elevated relative to the national average. The waterfall plot also reveals considerable geographic variation. Puerto Rico and the South US consistently showed higher meperidine usage than other areas. States in these geographic areas included the 1st, 2nd, 3rd, 4th, and 7th, highest areas in meperidine usage. There was a 32.8 fold difference in meperidine distribution in 2023 between the highest (Puerto Rico) and lowest (New Hampshire) areas (Figure 2). A heat map of the US and Puerto Rico visualizing meperidine usage in 2023 also shows this regional variation (Figure 3). The percent reduction in mean and median meperidine use between 2019 and 2023 was −62.5% and −63.3%. Significance was found for New Hampshire (−90.2%) and Alaska (−21.8%) (Figure 4).

**Fig 1.**
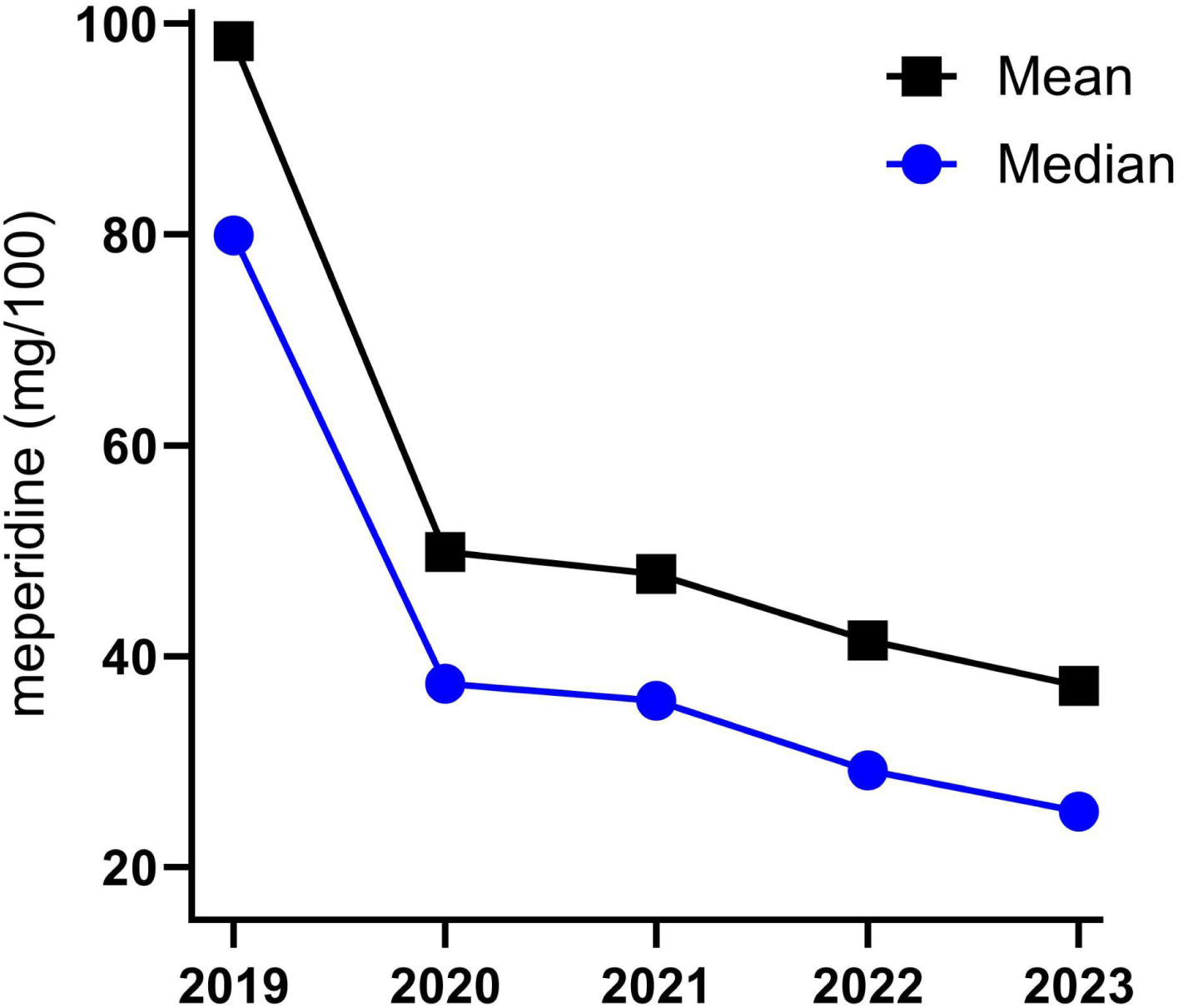
Meperidine mean and median distribution as reported to the US Drug Enforcement Administration’s Automated Reports and Consolidated Order Systems between 2019 and 2023.

**Fig 2.**
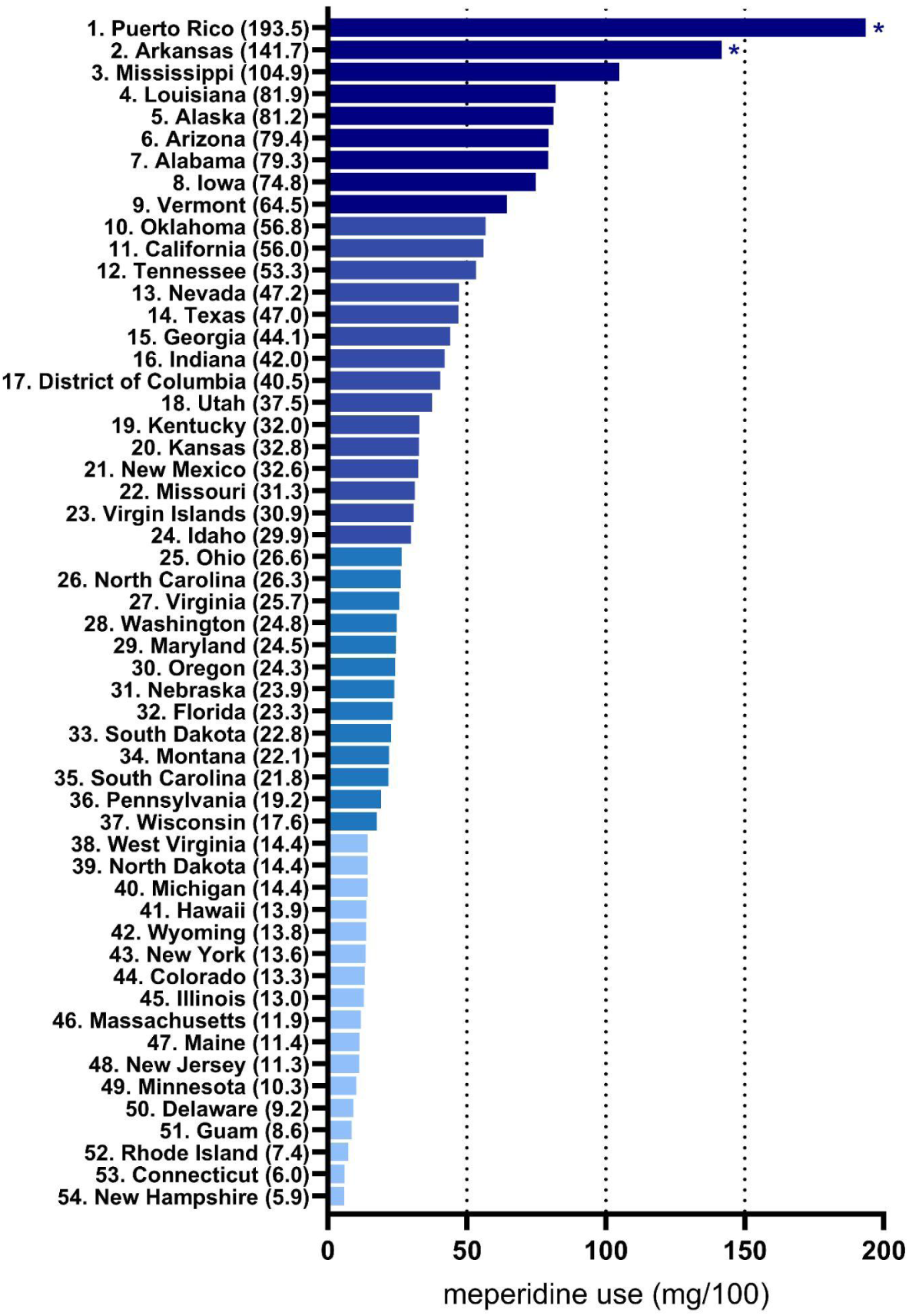
Ranked meperidine distribution as reported to the Drug Enforcement Administration’s Automated Reports and Consolidated Orders System in 2023 *p < .05 versus the national average.

**Fig 3.**
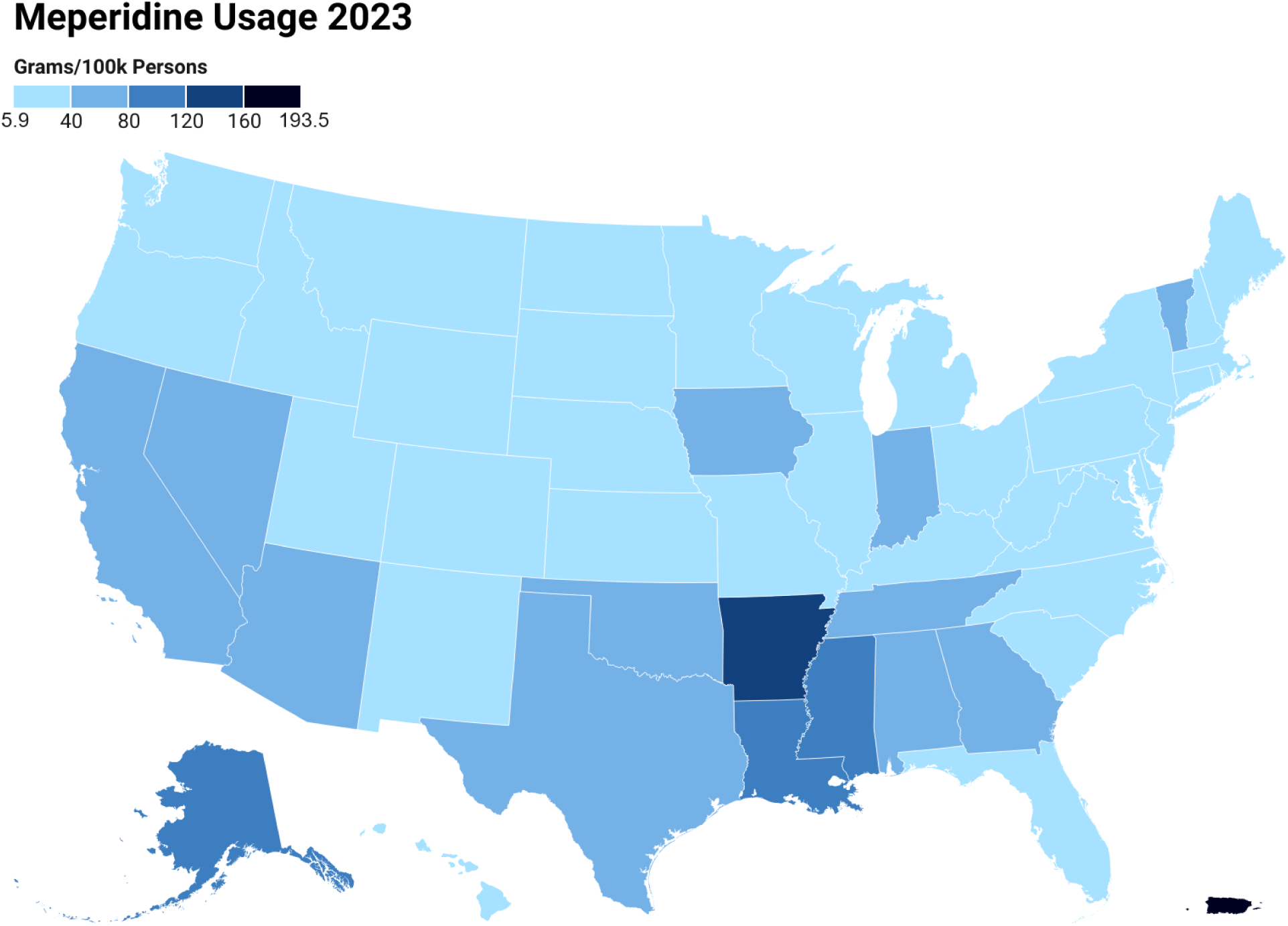
Heat map showing pronounced (32.8x fold) state level disparities in meperidine distribution as reported to the Drug Enforcement Administration’s Automated Reports and Consolidated Orders Systems in 2023.

**Fig 4.**
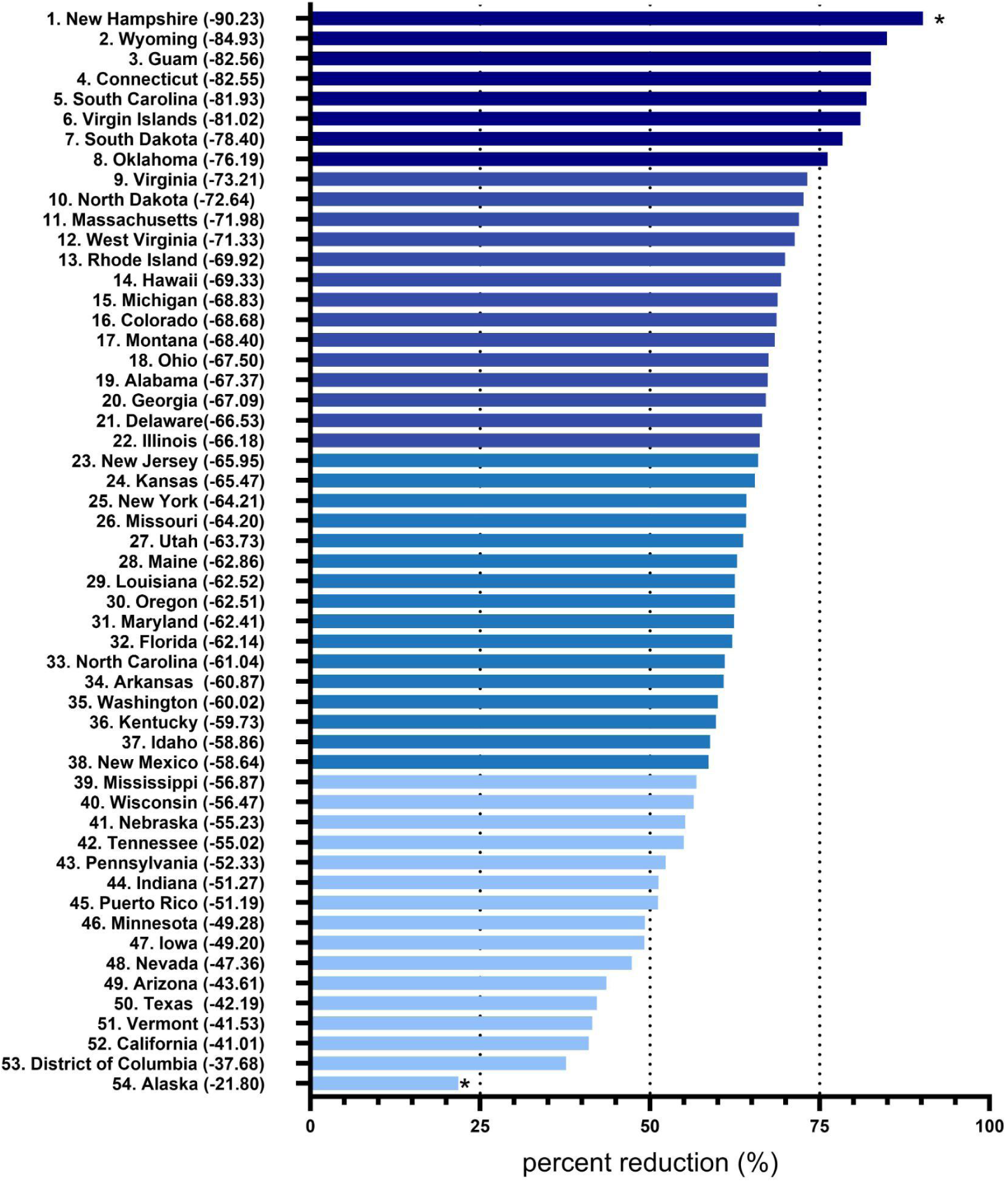
Percent decline in meperidine distribution as reported to the Drug Enforcement Administration’s Automated Reports and Consolidated Orders System from 2019 to 2023. *p < .05 versus the average (−62.48).

The meperidine distribution in 2023 by category for Arkansas, Puerto Rico, and Mississippi showed differences amongst areas. Puerto Rico and Mississippi had the majority of distributed meperidine by hospitals, with 68.91% and 62.10% respectively. However, the majority of Arkansas’s meperidine distribution was by pharmacies, with 57.32%. For all three areas, practitioners were linked with the lowest amount of meperidine distribution (Figure 5).

**Fig 5.**
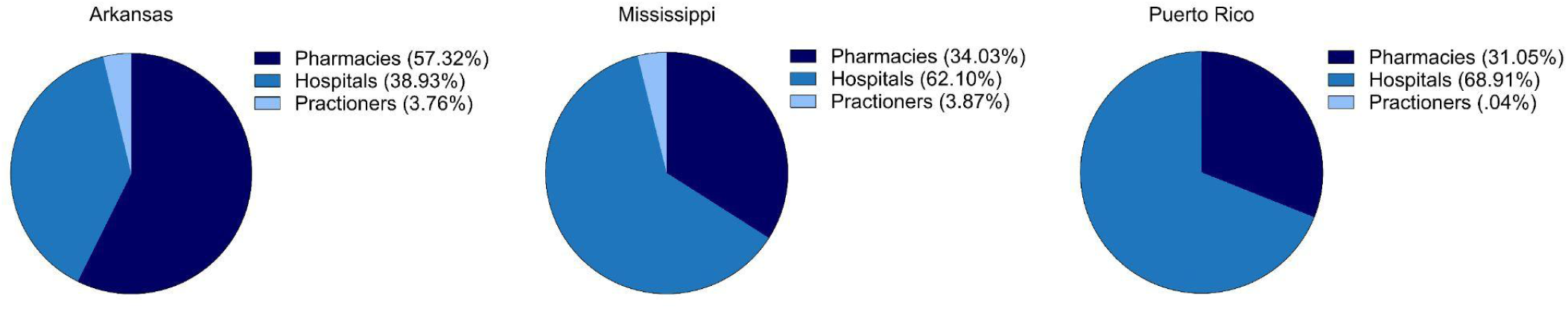
Pie charts showing meperidine distribution as reported to the Drug Enforcement Administration’s Automated Reports and Consolidated Orders System in 2023 by business activity.

Meperidine distribution for the first half of 2024 showed similar results to the pattern from 2019-2023. Puerto Rico (114.1) and Arkansas (64.2) had significantly more meperidine distribution than the rest of the US. Furthermore, states in the US South consistently had more usage than other regions, accounting for the states with the 2nd, 3rd, and 4th highest distribution, only behind Puerto Rico.

A linear regression between meperidine usage (mg/100 persons) and median household income in 2022 revealed significance (r(49) = -.38, p < .005). States in the South consistently showed lower relative incomes accompanied by higher meperidine usage (Figure 6). The linear regression between meperidine usage and median household income in 2023 had similar results (r(49) = -.58, p < .0001). Yet again, states in the South had lower relative incomes coupled with higher meperidine distribution (Figure 7). Lastly, linear regressions between meperidine usage (mg/100) and adult obesity percentage by state in 2022 showed a trend (r(52) = + .24, p = .078, Figure 8).

**Fig 6.**
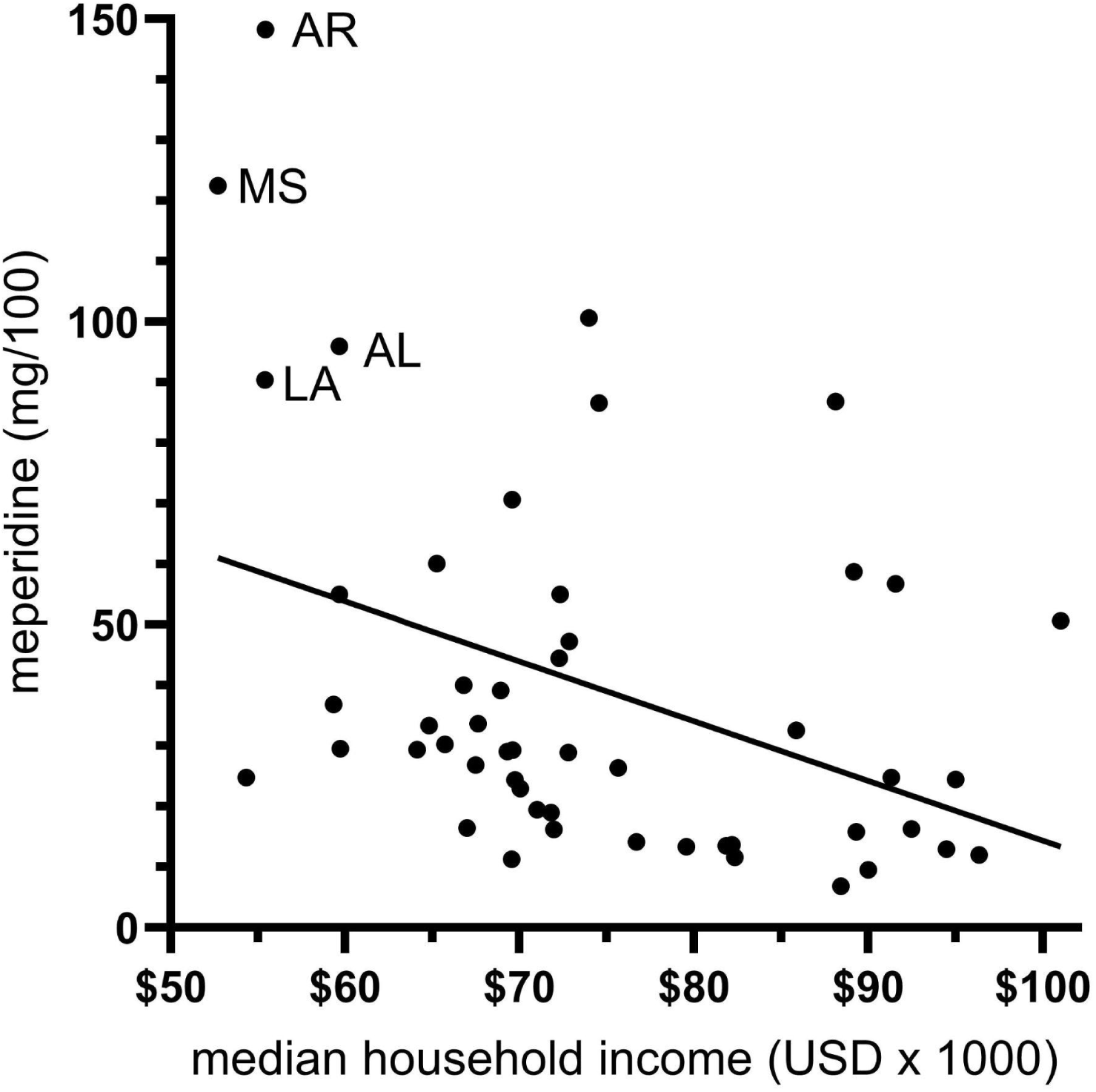
Significant (r(49) = -.38, p < .005) inverse association between median household income and meperidine distribution amongst the US states as reported by the Drug Enforcement Administration’s Automated Reports and Consolidated Orders System in 2022.

**Fig 7.**
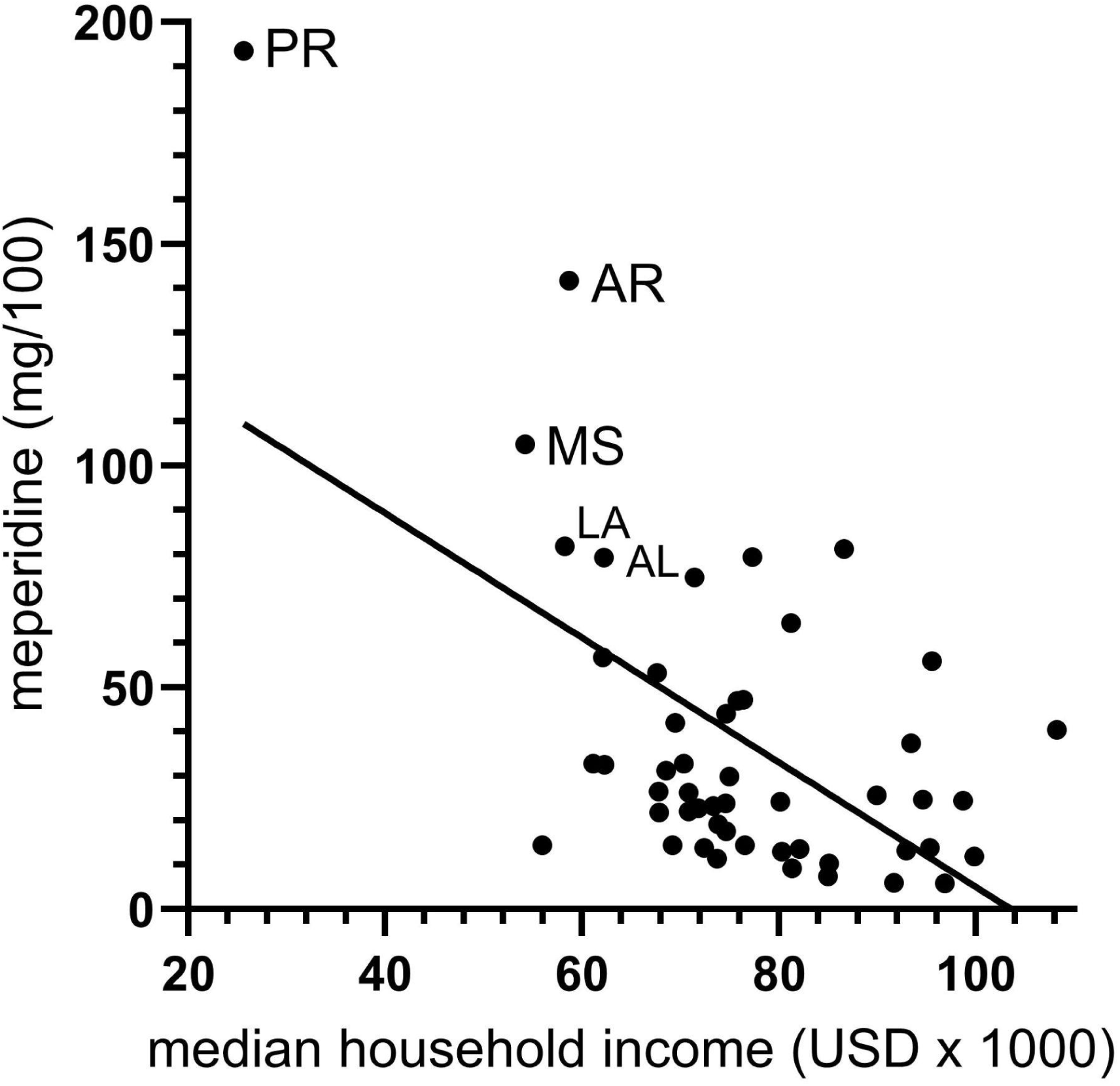
Significant (r(49) = -.58, p < .0001) inverse association between median household income and meperidine distribution amongst the US states as reported by the Drug Enforcement Administration’s Automated Reports and Consolidated Orders System in 2023.

**Fig. 8.**
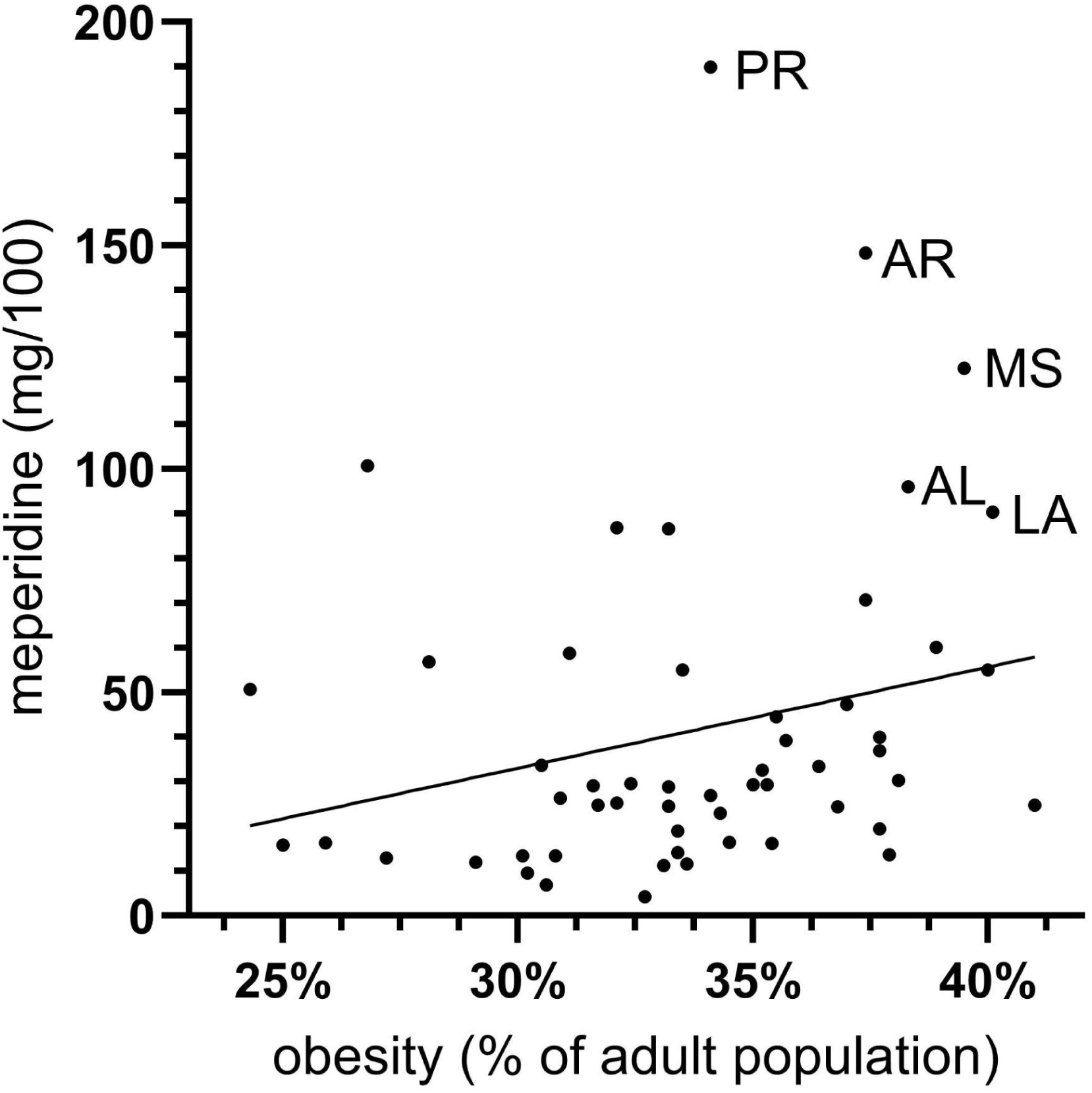
Linear regression between adult obesity prevalence and meperidine distribution in the US as reported by the Drug Enforcement Administration’s Automated Reports and Consolidated Orders System in 2022. (r(52) = .24, p = .078)

## DISCUSSION

The key finding from this pharmacoepidemiological study was that between 2019 to 2023, the total distribution of meperidine in the fifty states, Guam, the Virgin Islands, and Puerto Rico dropped by 57.8%. However, the majority of this decline occurred between 2019 and 2020, with a 48.3% reduction. From 2020 to 2023, there was only a decline of 18.5%. Prior reports found that meperidine distribution between 2000 and 2019 showed a fairly uniform reduction.^3^ Hence, this relative stagnation from 2020 to 2023 is of interest. Further research into why the reduction of meperidine distribution has plateaued may offer insights into possible strategies to decrease meperidine distribution more quickly as the years proceed. The use of educational interventions with support of other policy changes, such as formulary restrictions, has shown an ability to reduce meperidine distribution.^18^. Further analysis into meperidine education for healthcare professionals between the years 2019 to 2023 may shed light on the situation. However, this is only one possible answer, as health policy, drug costs, and other factors may have influenced the stabilization of meperidine reduction. The regional variation in meperidine distribution found by past studies persisted in this report.^3,4,19^ In 2019, the five states with the highest meperidine distribution were all in the South: Arkansas, Alabama, Mississippi, Oklahoma and Louisiana.^3^ Similarly, in 2020, the four states with the highest meperidine distribution were also all in the South: Alabama, Arkansas, Mississippi and Louisiana.^4^ This study identified a continuation of this regional variation. In 2023, when excluding Puerto Rico, the three states with the highest meperidine distribution were the Southern states: Arkansas, Mississippi, and Louisiana. In contrast, states in the Northeast in all three studies consistently had the lowest meperidine distribution.^3,4^ In this investigation, the three states with the lowest meperidine distribution in 2023 were: Rhode Island, Connecticut, and New Hampshire.

A recent study which investigated disparities in opioid distribution in Puerto Rico and the US between 2018-2023 was extended upon by our study.^19^ In the first half of 2024, Puerto Rico, Arkansas, Mississippi, and Lousiaiana led in meperidine distribution, echoing patterns of the past. Puerto Rico and Arkansas had significantly more meperidine distribution than the rest of the regions as reported by ARCOS as seen in supplemental figure 1.

We investigated these substantial regional variations amongst US states by analyzing possible associations between meperidine distribution, annual income, and adult obesity prevalence. A moderate inverse relationship was found between median household income and meperidine distribution in 2022 (r = −0.39, p < .005). Interestingly, Southern states, including Arkansas, Mississippi, Louisiana, and Alabama all populated the upper left quadrant of the scatter plot. A stronger inverse relationship was found in 2023 (r = -.58, p < .0001). Furthermore, Southern states remained in the upper left quadrant of the scatter plot. Median income was used as a variable in this study in order to introduce the broader topic of affordability in the current healthcare system. This significant correlation opens the door to future research into the affordability of better opioid options in states with lower median incomes. Other research opportunities can look into the common insurances and benefits found within states with lower median incomes such as the Southern states or Puerto Rico.

A positive trend was found between adult obesity prevalence and meperidine distribution in 2022 (r = .242, p = .078) (Figure 8). A prior study also identified this relationship for the year 2019.^3^ According to the earlier report, the connection between the two variables was acute pancreatitis, a health issue that can be treated with meperidine and caused by obesity.^3^ Yet again, states located in the south were clustered in the upper left quadrant, signifying higher rates of adult obesity and meperidine distribution.

The use of ARCOS in pharmacoepidemiologic research has both strengths and weaknesses. A particular asset of the database is that it is comprehensive for meperidine and other Schedule II Substances distributed in both states and US territories. That being said, the database does not differentiate between formulations. Furthermore, continued research with other databases will be needed to identify specific health professions that continue to use meperidine.

In conclusion, while meperidine distribution has continued to decline between 2019 and 2023, the decline has largely stagnated. This was accompanied by the persistence of substantial state-level variations. Significant associations were found between meperidine distribution, median income, and adult obesity prevalence. Further research into other possible correlates such as drug costs and insurances by location can shed more light into regional variations of meperidine usage. These findings, and further research, also have the potential to influence public policy. Whether it be incentives to use safer alternatives or the development of better alternatives, research in this field has the potential for public change. Altogether, the known risks associated with meperidine usage, and access to alternative agents, should, and has, played a role in the continued decline.

## ACKNOWLEDGEMENTS

The Geisinger College of Health and Science’s Center of Excellence Program and the Pathways Program supported this research.

## FUNDING

Geisinger College of Health and Science Summer Research Immersion Program.

## DISCLAIMER

All authors write with individuality and do not necessarily represent the views of their affiliations.

## COMPETING INTERESTS

Brian J. Piper was (2019-2021) a member of an osteoarthritis research team that was supported by Eli Lilly and Pfizer. All other authors have no competing interests.

## PATIENT CONSENT FOR PUBLICATION

Not required.

## ETHICS APPROVAL

Approved by Geisinger as Exempt.

## DATA AVAILABILITY STATEMENT

All data analyzed in this study is available through the following public archives: ARCOS,^9^ US Census Bureau,^10–12^, ^14^ UN,^13^ and CDC.^15^

---

**Supplemental Figure 1.**
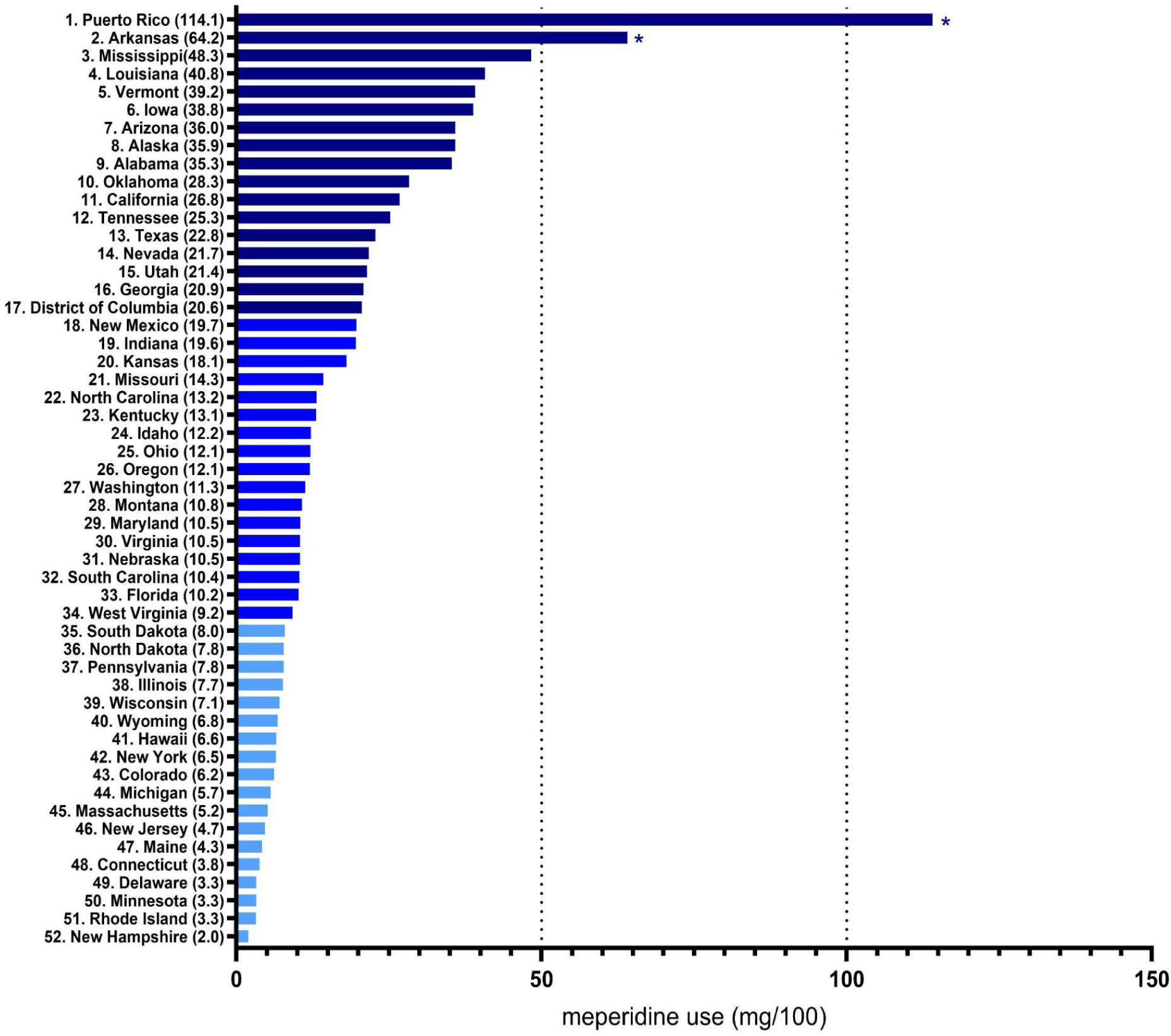
Ranked meperidine distribution per 100 persons as reported to the US Drug Enforcement Administration’s Automated Reports and Consolidated Orders System in 2024. *p < .05 versus the national average.

**Supplemental Figure 2.**
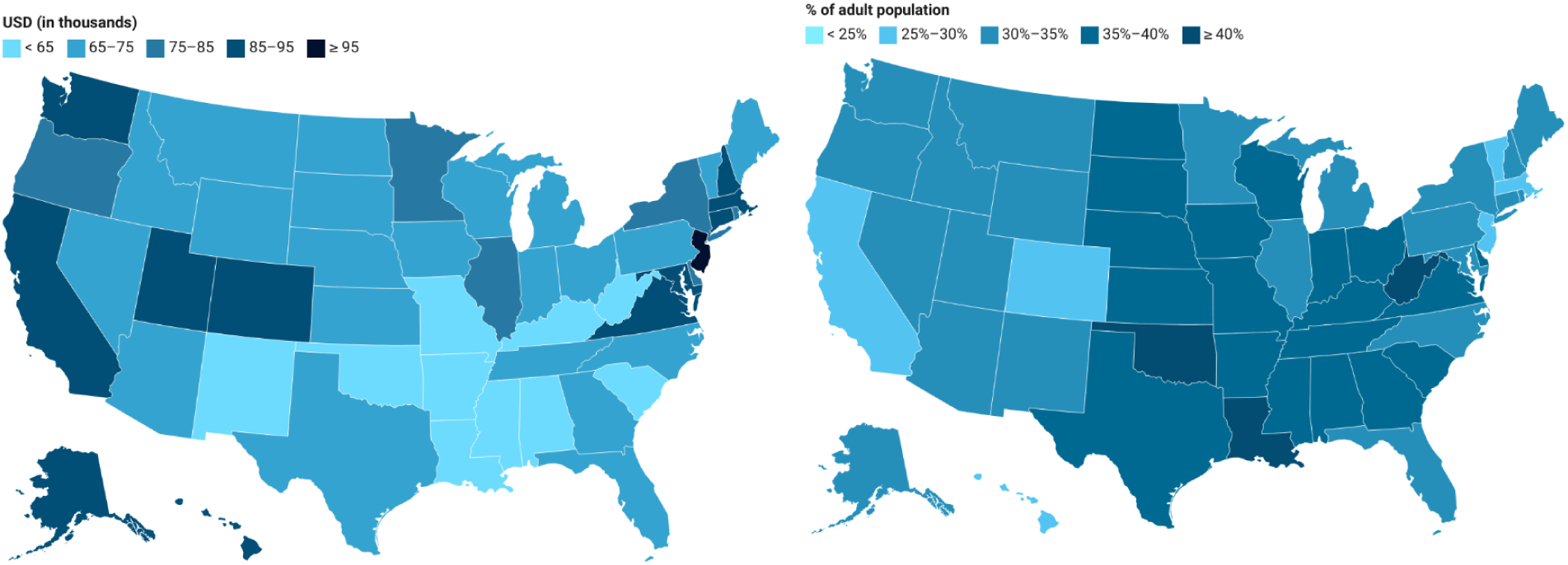
Heat maps of median household income and percent adult obesity in 2022.

## Notes

### Funding Statement

This study was funded by the Geisinger College of Health and Sciences Summer Research Immersion Program.

### Author Declarations

All data analyzed in this study is available through the following public archives: ARCOS, US Census Bureau, and the CDC.

